# A deductive approach to modeling the spread of COVID-19

**DOI:** 10.1101/2020.03.26.20044651

**Authors:** Pranav Mishra, Shekhar Mishra

## Abstract

Severe acute respiratory syndrome coronavirus 2 (SARS-COV-2), previously known as 2019-nCoV, is responsible for the atypical pneumonia pandemic designated as Coronavirus Disease 2019 (COVID-19). The number of cases continues to grow exponentially reaching 492,000 people in 175 countries as of March 25, 2020. 22,169 people (∼4.5%) infected with SARS-COV-2 virus have died. We have developed an exponential regression model using the COVID-19 case data (Jan 22 – Mar 22, 2020). Our primary model uses designated *Phase 1* countries, who exceed 2500 cases on Mar 22. The model is then applied to *Phase 2 countries*: those that escaped the initial *Phase 1* global expansion of COVID-19. With the exception of *stabilizing countries* (South Korea, Japan, and Iran) all *Phase 1 countries* are growing exponentially, as per *I*_2500_(*t*) = 120.4 × *e*^0.238*t*^, with a rate, *r* = 0.238 ± 0.068. Excluding China, the BRICS developing nations and Australia are in *Phase 2*. Case data from *Phase 2 countries* are following the model derived from *Phase 1 countries*. In the absence of measures employed to flatten the curve including social distancing, quarantine, and healthcare expansion, our model projects over 274,000 cases and 12,300 deaths in the US by Mar 31. India can expect 123,000 cases by April 16. By flattening the curve to the growth rate of *stabilizing countries* (*r* = 0.044 ± 0.062), the US would prevent 8,500 deaths by Mar 31, and India would prevent 5,500 deaths by April 16.

## Introduction

Severe acute respiratory syndrome coronavirus 2 (SARS-COV-2), previously known as 2019-nCoV, is a novel virus in the coronaviridae family of positive sense, enveloped, RNA viruses^1^. It is responsible for the atypical pneumonia pandemic designated as Coronavirus Disease 2019 (COVID-19), by the World Health Organization. First identified in Wuhan, China in December 2019, COVID-19 has spread to 175 countries/territories, with over 492,000 cases as of March 25, 2020^2,3^. During this period, there has been a stepwise escalation of international, national, and regional governmental responses. The Wuhan Government confirmed that local hospitals were treating 27 cases of viral pneumonia on December 31, 2019^4^. However, approximately three weeks later, on January 20, the United States confirmed its first case^5^. Due to the rapid expansion of cases across the world, the World Health Organization (WHO) designated COVID-19 as Public Health Emergency of International Concern on January 30, 2020^6^. On the same day, India confirmed its first case in the state of Kerala^7^.

The United States Centers for Disease Control and Prevention (CDC) issuing a travel alert to the Wuhan region on January 6, 2020^8^. Various countries systematically blocked international travel from known COVID-19 hot-spots, including China, Iran, and the European Union. At the time of writing, nearly 80 nations have a global travel ban with an additional 9 implementing global quarantine measures^9^. These efforts hope to prevent local seeding of infection, each of which can rapidly increase case numbers. When examining smaller confined communities exposed to SARS-COV-2, the disease spreads unabated. Initial studies examined high-density exposed cohorts, including cruise ships, prisons, nursing homes, and healthcare workers^10–13^. Therefore, it appears that containment of COVID-19 primarily be mitigated by preventing local seeding through travel restriction and reducing the population density around potential infected person.

A wide array of models exists for the predicting the spread of infectious disease across various cohorts. These models may be stochastic in nature, creating probability distributions by moving about random variables, or deterministic in nature, compartmentalizing the population across various groups. The Susceptible Exposed Infectious Recovered (SEIR) model is a widely utilized mathematical model in the COVID-19 outbreak^14^. Liu et al reviewed the models of 12 studies which calculated the basic reproductive number (R_0_) of the SARS-COV-2 virus^15^. The studies examined employed various techniques of subdividing the population and utilizing historical data of related diseases (i.e. SARS, MERS) to estimate the potential impact of COVID-19. While these models help simulate the possible course of a disease given population and disease characteristics, each model becomes outdated as various governments implement restrictions on its population. Infectious disease dynamics are classically considered as exponential in nature. However, Maier and Brockmann demonstrate that COVID-19 is affected by “fundamental mechanisms that are not captured by standard epidemiological models” ^16^. This is notably true in early phases of disease expansion. Consequentially, COVID-19’s behavior warrants the examination of models outside of inductive logic.

The authors of this paper seek to understand the expansion of COVID-19 through a deductive examination of existing case data. Our study examines this pandemic through data provided by the Johns Hopkins University’s Systems Science and Engineering Group (JHU CSSE)^17^. Our approach is inherently simplistic as it examines COVID-19 case numbers first. This deductive ‘outside-in’ observational approach allows us to derive models without consideration of ever-changing population dynamics reflective of health policy measures being implemented. We seek to apply our model to nations which have avoided the initial expansion of COVID-19. In this study, we model the pandemic’s impact on the United States and BRICS developing nations. India’s 657 cases, as of March 25, represent ample seeding, priming the nation for a devastating expansion of cases and fatalities.

## Methods and Results

### Raw Data - COVID-19 Case Numbers

The pandemic case data is provided by JSU CSSE’s GitHub repository on COVID-19. It is an aggregation of case data starting on January 22, 2020, from several sources^17^. Cases in the data set include laboratory confirmed cases and presumptive cases of COVID-19.

#### Sampled Data

- *Phase 1 Countries*: All countries reporting greater than 2500 cases as of March 22, 2020
- *Members of the Group of 7* (*G7*): Canada, France, Germany, Italy, Japan, UK, US
- *Members of the BRICS developing nations*: Brazil, Russia, India, China, South Africa
- *Developed nations of interest in Phase 2*: Australia

We include a series of countries into our data set for comparative purposes. We designate *Phase 1 countries* as those who exceed 2500 cases of COVID-19 as of March 23, 2020. These countries, which suffered the initial global expansion of SARS-COV-2, provide a working example for countries who recently acquired the virus. We also include members of the Group of 7 (G7) to examine differences between developed and *Phase 1* countries. We include BRICS countries as an example of large, developing countries likely to become part of *Phase 2*. The *Phase 2* countries are hypothesized to acquire SARS-COV-2 either through direct seeding from the epicenter, China, or via secondary seeding from *Phase 1* countries. We include Australia as a nation of interest, due to its developed status, geographic proximity to China, and relative escape from *Phase 1*.

### Excluding Data Prior to the 100^th^ Case Per Country

In the early expansion of COVID-19 within each country, cases primarily expanded from international seeding from China, rather than internal human-to-human spread. The sporadic nature of these cases spreading around the world can be visualized through notable sub-exponential, erratic growth patterns, within a country. After a country reaches its 100^th^ case, the disease load increases exponentially (**Figure 1**). The time it takes to exceed 100 cases also demonstrates substantial variation (**Figure 2**).

**Figure 1:**
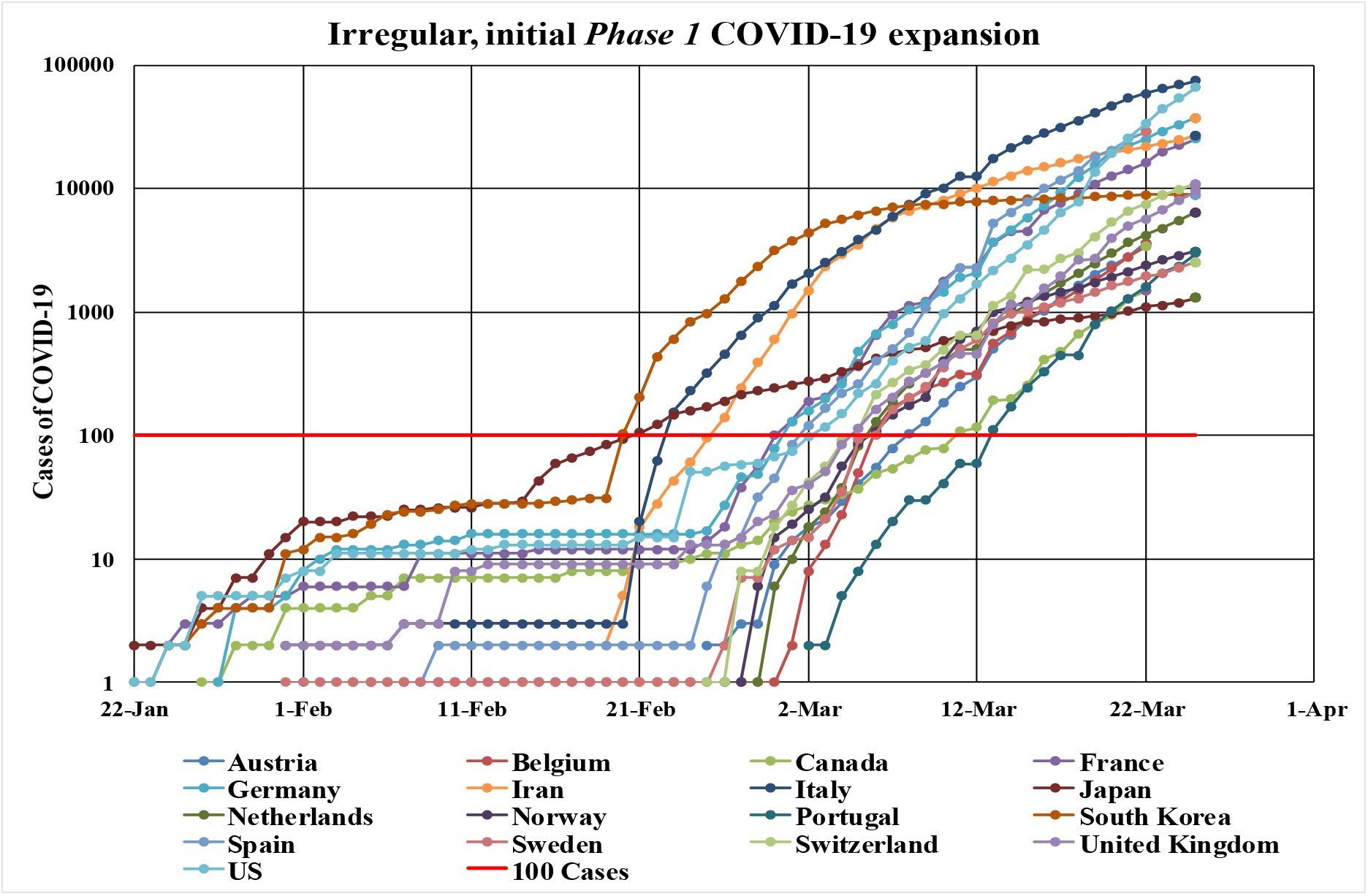
Irregular, initial Phase 1 COVID-19 expansion. Total number of COVID-19 cases in countries reporting greater than 2500 cases, as of March 23, 2020. JHU CSSE data collection on COVID-19 cases starts Jan 22, 2020. A red line is placed indicating 100 cases. Below the line, we note sub-exponential, erratic growth patterns. During this time, cases primarily expand from international seeding, rather than internal human-to-human spread. Therefore, we exclude data below 100 cases, per country.

**Figure 2:**
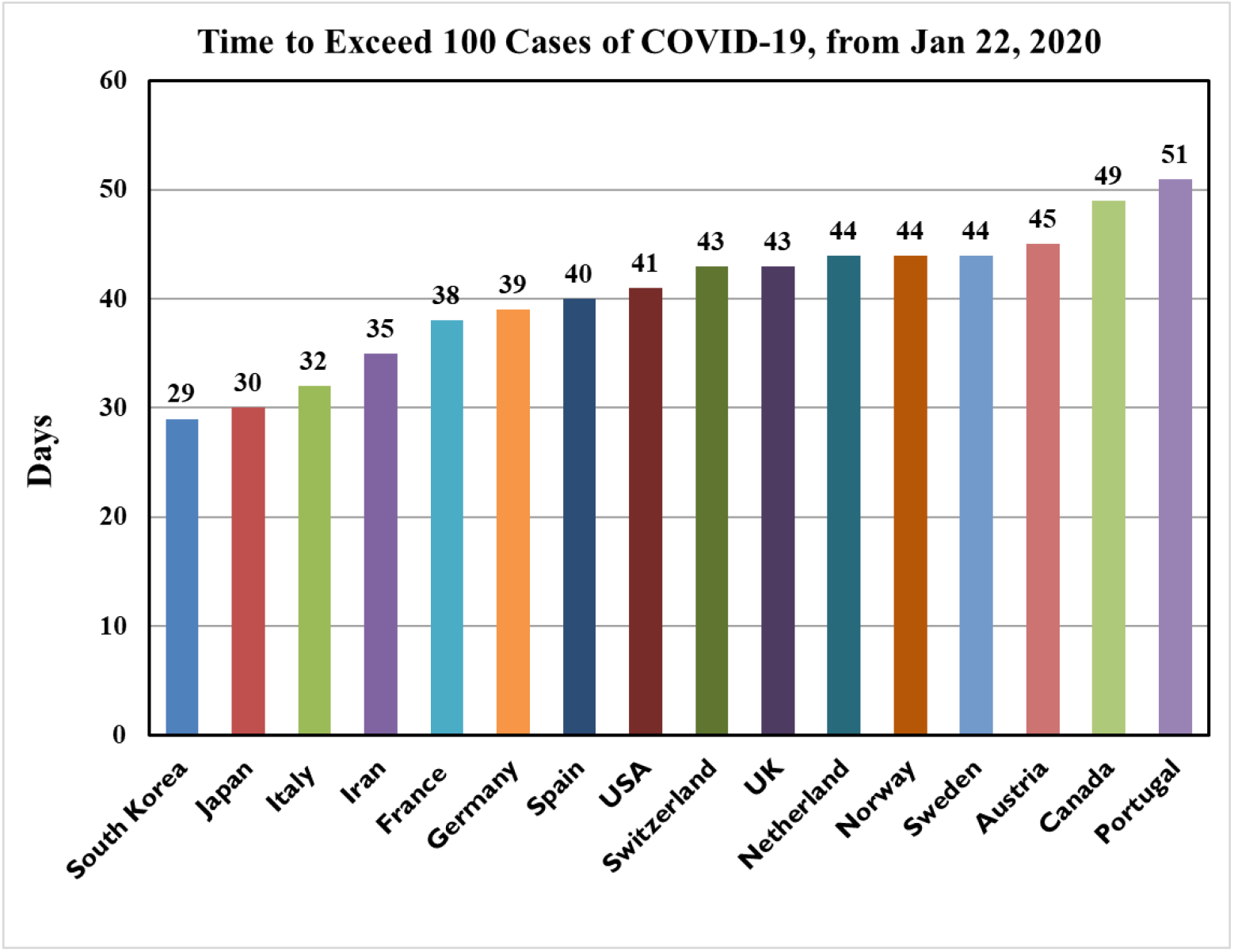
Days to exceed 100 cases of COVID-19 from January 22, 2020. The JHU CSSE data set starts on January 22, 2020. From this date, we count the number of days each country takes to exceed 100 COVID-19 cases. The initial expansion is primarily from international seeding, rather than internal human-to-human spread. Countries which reached 100 cases likely have greater international travel with China. For the countries shown, the number of days to reach 100 cases has a mean of 40 days, with standard deviation 6.4 days.

Of the countries already exceeding 2500 cases, the mean time to exceed 100 cases is 40 days, with standard deviation 6.3 days, calculated from Jan 22, 2020. The high variance in data supports excluding the first 100 cases from the modeling equations.

### Inclusion Criteria for the Predictive Model

- Model 1: All countries reporting greater than 2500 cases as of March 23, 2020
- Model 2: G7 countries

### Exclusion Criteria for the Predictive Model

We exclude four countries: China, Iran, Japan, and South Korea. China is excluded due to notably unreliable data and changed its methodology. Numerous reports exist demonstrating the suppression of case data by Chinese authorities^18–20^. Iran, Japan, and South Korea are excluded as outliers, which approached the stationary phase of COVID-19 early in the timeline (**Figure 3**).

**Figure 3:**
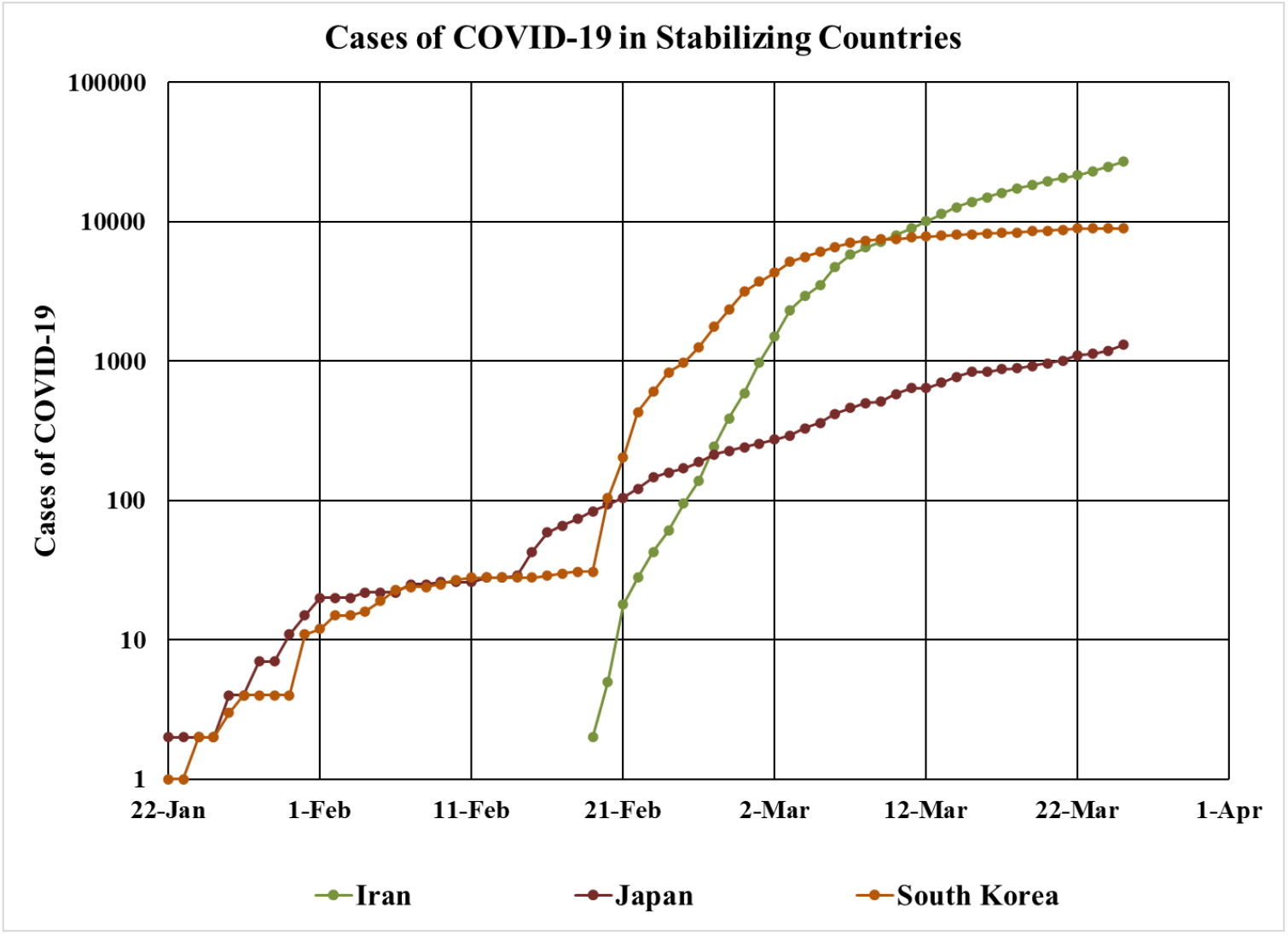
Cases of COVID-19 in Stabilizing Countries. We isolate Iran, Japan, and South Korea from Fig. 1. On a logarithmic scale graph, the rate of exponential growth is visualized by the slope of the curve. These countries obtained early COVID-19 expansion reduction, noted by a significant change in slope. They are excluded from the modeling equations.

### Creating a Predictive Model

Eleven countries pass the inclusion and exclusion criteria. For each of them, an exponential regression is applied, as per the function:

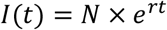

where *I* is the number of infected cases at a time t in days, *N* is the initial case load, *r* is the rate of growth. The results are reported in **Tables 1-3*Table 1***. We then average the values for *N* and *r*, producing:

**Table 1:**
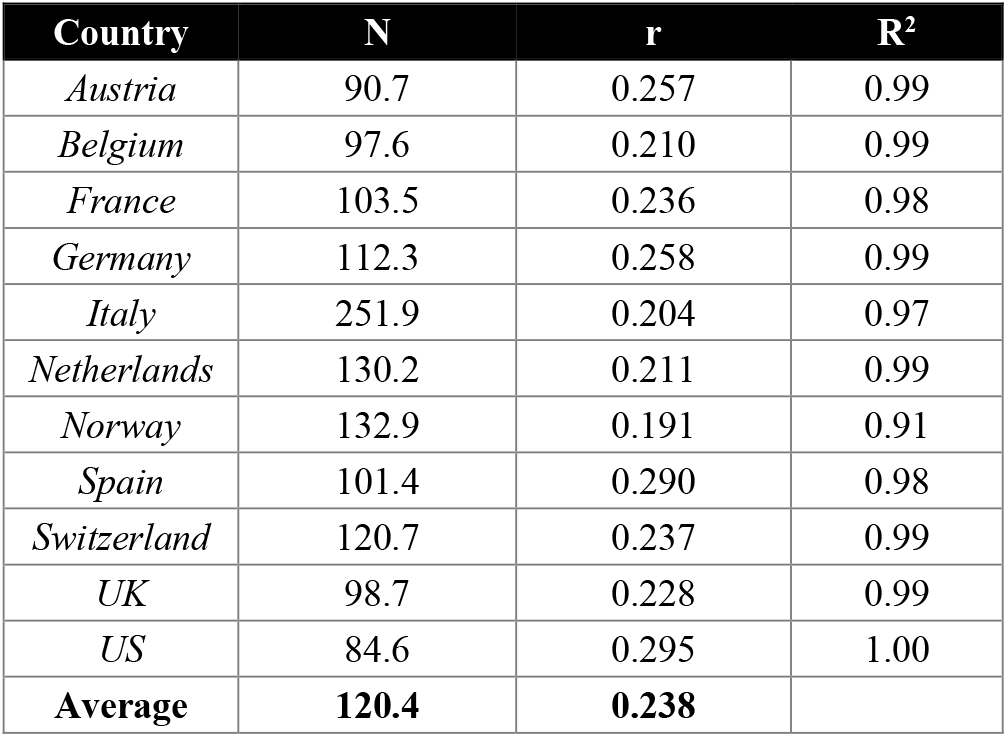
Exponential regression modeling of Phase 1 countries with greater than 2500 cases as of March 22, 2020, for the equation I(t) = N×e^rt^. R^2^ correlation coefficients are included per country. An average value for ‘N’ and ‘r’ is calculated. The mean rate of expansion of Phase 1 countries is 0.238, with standard deviation 0.034.

Analysis
| Country | N | r | R<sup>2</sup> |
| --- | --- | --- | --- |
| *Austria* | 90.7 | 0.257 | 0.99 |
| *Belgium* | 97.6 | 0.210 | 0.99 |
| *France* | 103.5 | 0.236 | 0.98 |
| *Germany* | 112.3 | 0.258 | 0.99 |
| *Italy* | 251.9 | 0.204 | 0.97 |
| *Netherlands* | 130.2 | 0.211 | 0.99 |
| *Norway* | 132.9 | 0.191 | 0.91 |
| *Spain* | 101.4 | 0.290 | 0.98 |
| *Switzerland* | 120.7 | 0.237 | 0.99 |
| *UK* | 98.7 | 0.228 | 0.99 |
| *US* | 84.6 | 0.295 | 1.00 |
| **Average** | **120.4** | **0.238** |  |

**Table 2:** Exponential regression modeling of Group of 7 (G7) countries, for the equation I(t) = N×e^rt^. An average value for ‘N’ and ‘r’ is calculated. The mean rate of expansion is 0.245, with standard deviation 0.0305. Japan, a member of the G7, is displayed as a strikeout due to being present in the exclusion criteria. However, we display it for comparison purposes. Note that Japan’s rate of expansion, r, is approximately 3 times below the average for G7 nations.

| Country | N | r | R<sup>2</sup> |
| --- | --- | --- | --- |
| *Canada* | 81.0 | 0.249 | 0.99 |
| *France* | 103.5 | 0.236 | 0.98 |
| *Germany* | 112.3 | 0.258 | 0.99 |
| *Italy* | 251.9 | 0.204 | 0.97 |
| *Japan* | 119.6 | 0.077 | 0.98 |
| *UK* | 98.7 | 0.228 | 0.99 |
| *US* | 84.6 | 0.295 | 1.00 |
| **Average** | **122.0** | **0.245** |  |

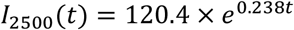

***Model Equation 1: Countries Exceeding 2500 cases of COVID-19***

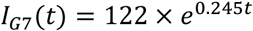

***Model Equation 2: Group of 7* (*G7*) *Countries***. *Japan is excluded from the model due as it meets the exclusion criteria*.

**Figure 4** presents our primary model, I2500, with 95% confidence intervals and case data of Phase 1 countries. We aligned the data from each Phase 1 country, such that t = 1 day when the number of cases is closest to, but greater than 100. The 95% confidence intervals continue to contain the trajectories of nearly all Phase 1 countries beyond Mar 22.

**Figure 4:**
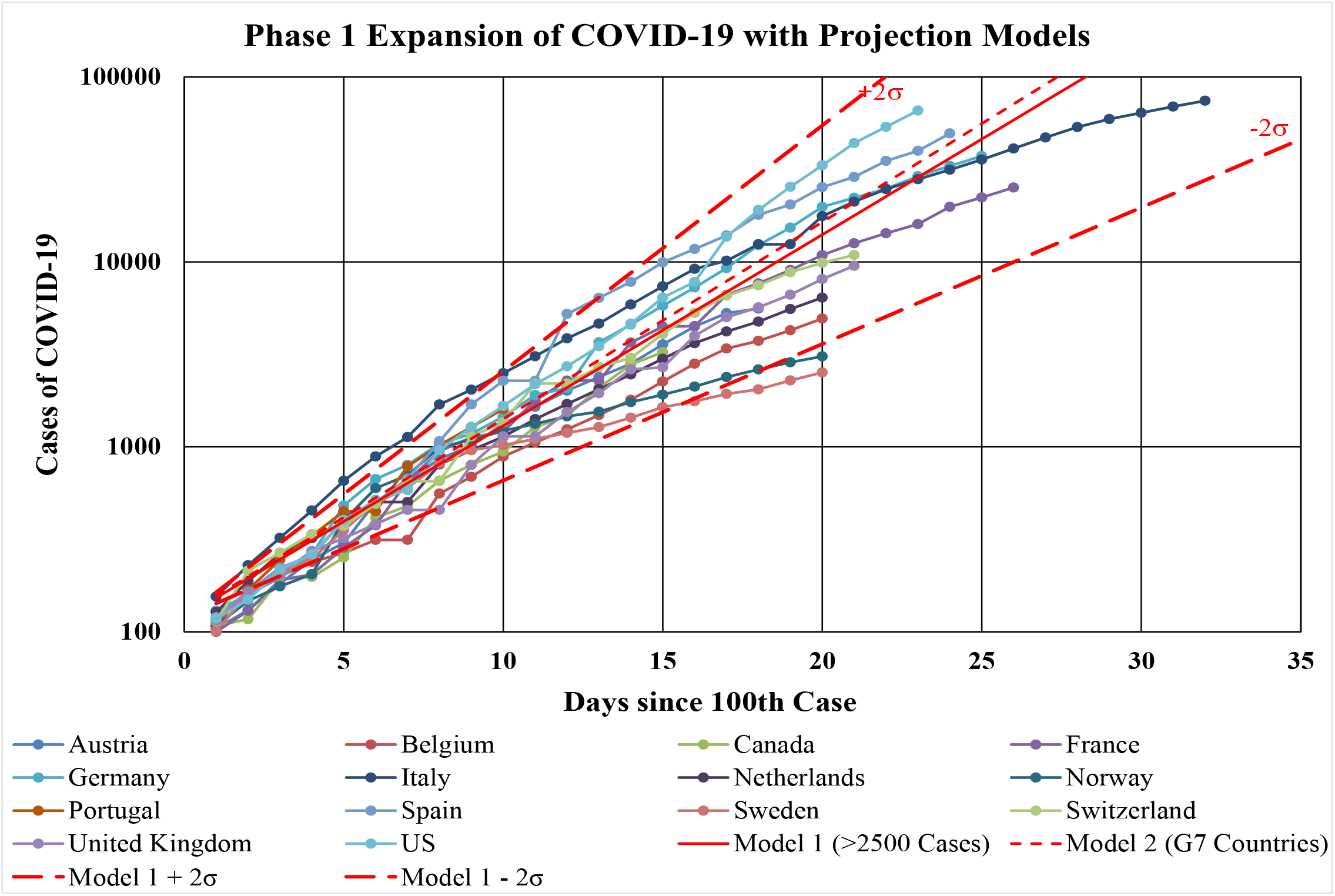
Phase 1 Expansion of COVID-19 with Models. We aligned the data from each Phase 1 country, such that t=1 day when the number of cases is closest to, but greater than 100. We include 95% confidence intervals in red, which contain the trajectory of most countries. Disparity is expected near t=1, as countries expand beyond the 100-case cutoff at varying rates. Many reasons exist for early variation in case detection, including, but not limited to, differences in international seeding from China, testing capabilities, healthcare systems, public health policy, including quarantine measures.

## Analysis

### Applying the Model to Developing Nations

We have excluded the SARS-COV-2 source nation of China from the BRICS countries for future modeling. We hypothesize that these major developing countries escaped the first phase of global COVID-19 expansion due to reduced seeding. With a smaller amount of air travel between China and the other BRICS countries, the time to reach the 100^th^ case will fall later in the pandemic’s timeline. Next, we fit the equation, *I*_2500_(*t*) = 120.4×*e*^0.238*t*^, to each country **(Figure 5)**. For *t=1 day*, the model projects 152 cases. Thus, we start the model for each country on the date with number of COVID-19 cases closest to 152.

**Figure 5:**
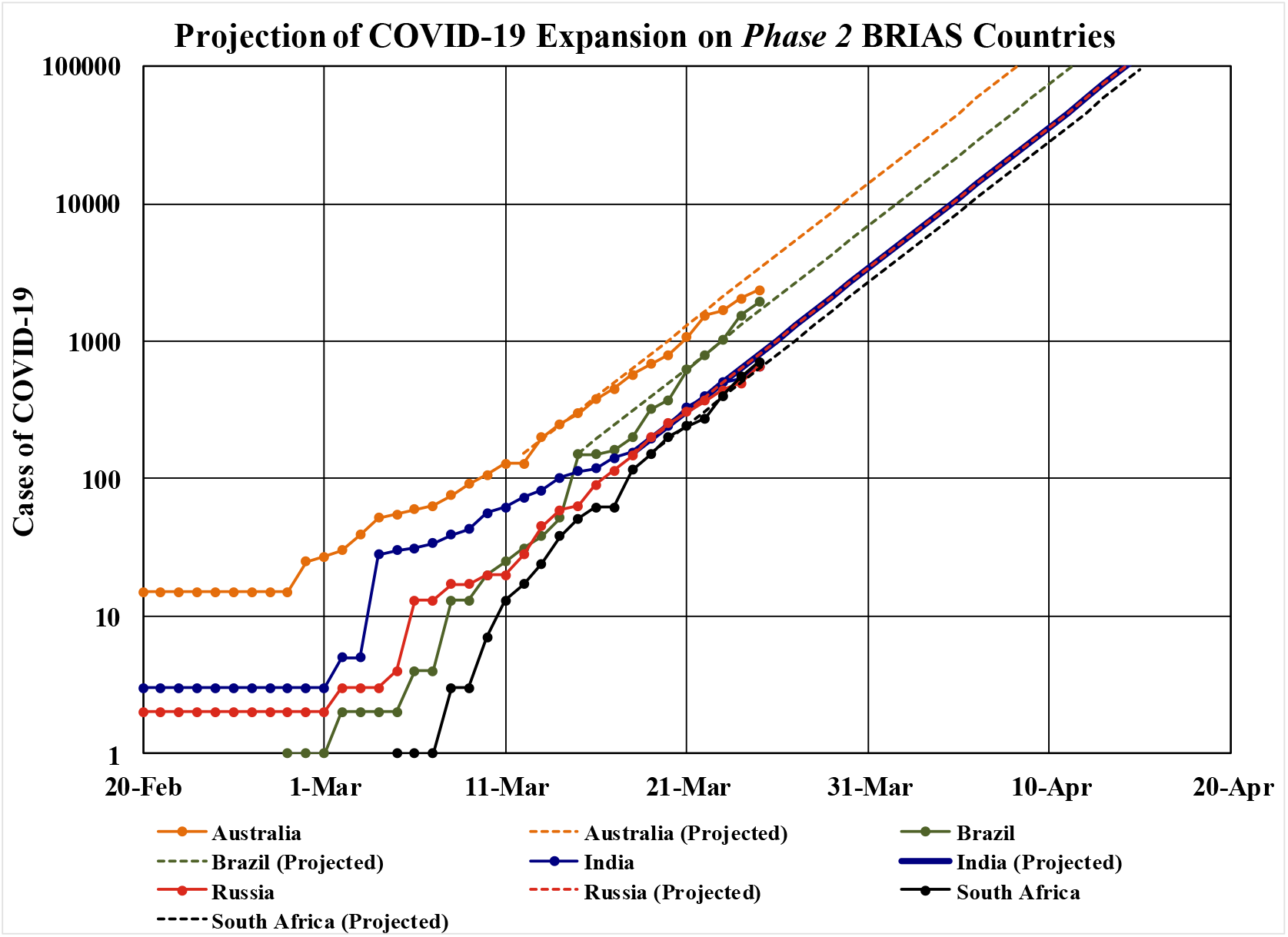
Projection of COVID-19 expansion on Phase 2 BRIAS countries. This group of countries includes the large developing nations of Brazil, Russia, India, and South Africa. We have additionally included Australia as a country of interest, which escaped the initial international expansion of COVID-19.

The actual number of infected persons will be affected by statistical uncertainties. We use our fit and standard deviation in the fitted parameter to estimate the range of growth at ± 1s and 2s levels for India. We expect all *Phase 2 countries* to have a similar band of cases (**Figure 6**).

**Figure 6:**
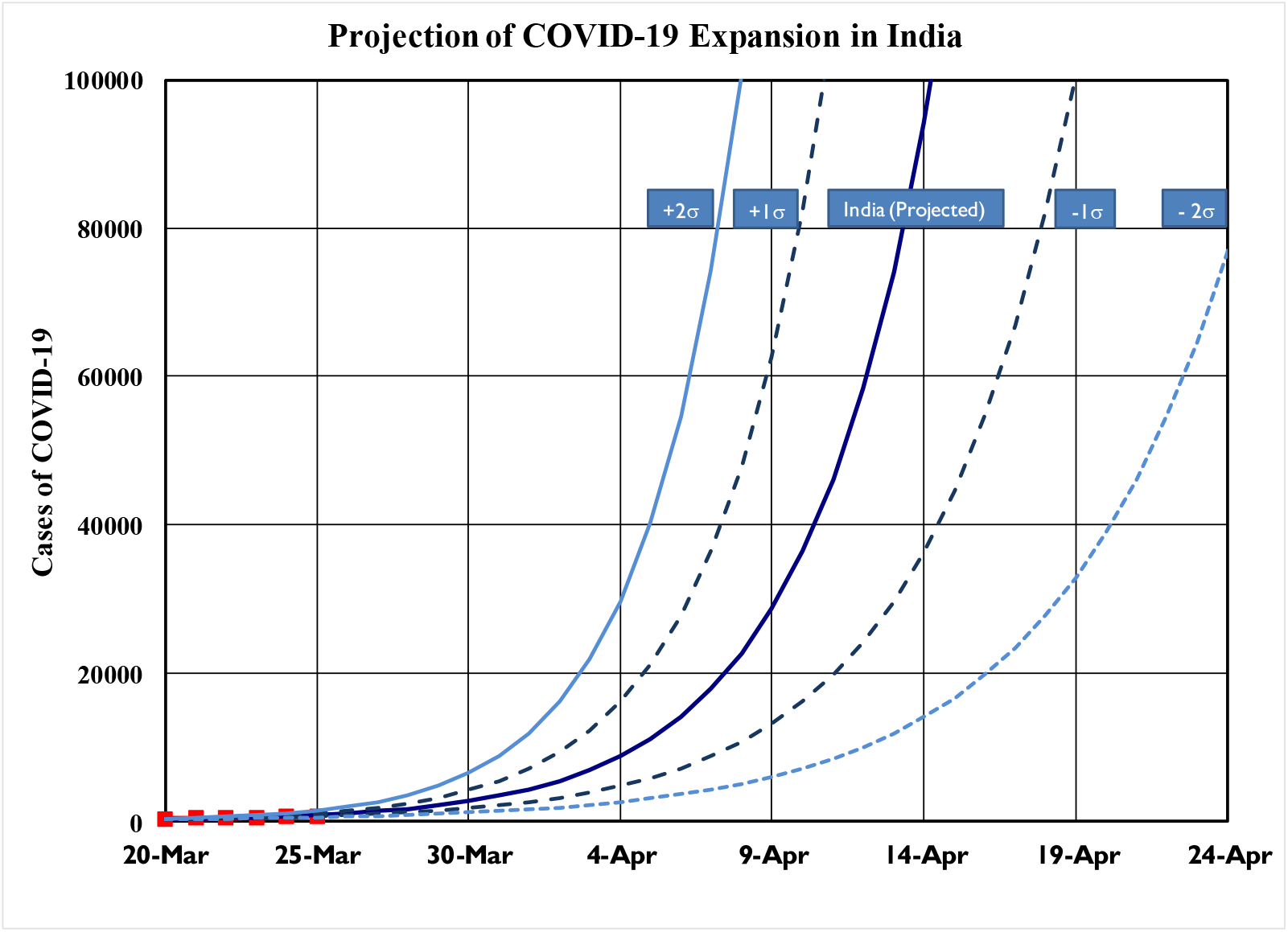
Projection of COVID-19 Expansion in India. The mean rate (r) of expansion of Phase 1 countries is 0.238, with standard deviation (σ) 0.0340. We plot 1s and 2s variations about the function I_2500_(t) = 120.4×e^0.238t^. There is a 95% probability that India’s case rate will fall between the outer two lines, assuming it COVID-19 expands at the same rate as Phase 1 countries. Existing case data is plotted with red markers.

### Modeling the Impact of Stabilizing Expansion

For theoretical consideration, we examine what would happen if a country could instantaneously shift from the *Phase 1* growth rate to that of the *stabilizing countries*, starting March 26, 2020.

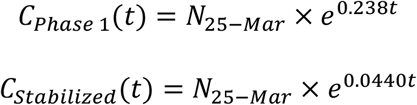

***Model Equations 3 and Model Equation 4:*** *N*_25−*Mar*_ *as the number of cases in a country on March 25 and t is the number of days after March 25. The growth rates are derived from Tables 1 and 3, respectively*.

**Table 3:** Exponential regression modeling of stabilized countries, from March 12-22, 2020, for the equation I(t) = N×e^rt^. An average ‘r’ is calculated at 0.044. This post-stabilized average rate of growth is 5.36 and 5.52 times smaller than the average rate of growth of Phase 1 and G7 countries, respectively.

| Country | r | R<sup>2</sup> |
| --- | --- | --- |
| *Iran* | 0.075 | 0.980 |
| *Japan* | 0.046 | 0.946 |
| *South Korea* | 0.012 | 0.991 |
| **Average** | **0.044** |  |

The estimated case fatality rate (CFR) of the COVID-19 is 4.51%, based on arithmetic division of fatalities to known cases^17^. We can then estimate the number of preventable deaths when if a country instantly stabilized its growth to that of South Korea, Japan, and Iran.

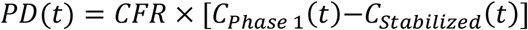

***Model Equation 5: Preventable deaths with Phase 1 growth***. *PD is preventable deaths. The case fatality ratio of COVID-19* (*CFR*) *is 4*.*51%. It is multiplied into the difference of cases between Phase 1 growth and that of stabilizing countries*.

## Discussion

Through our examination of COVID-19’s expansion around the world, we draw several conclusions on the intrinsic nature of SARS-COV-2 and the impact of human intervention to curtail its spread. First, we note that most countries affected in the *Phase 1* growth of disease follow exponential growth curves with comparable rates. Norway exhibits the minimum rate at 0.191, while the United States has the greatest at 0.295. The mean rate of expansion of *Phase 1* countries is 0.238 (95% CI 0.170-0.306). For G7 countries, the mean rate of expansion is 0.245 (95% CI 0.184-0.306). Thus, we observe that G7 nations follow closely with other *Phase 1* nations in COVID-19 cases.

When compared with the three stabilized countries of Iran, Japan, and South Korea, we notice more troubling picture. The average rate of growth of stabilized countries is in excess of 5 times less than the rate of growth of *Phase 1* and G7 countries. If the US follows *I*_*2500*_, we expect over 119,000 cases by Mar 31. More concerning, though, is that the US is growing one standard deviation above *I*_*2500*_. The cases based on *C*_*Phase*1_is over 274,000 by Mar 31. If it were possible to instantaneously change to the rate of growth calculated for stabilized countries, the expansion of COVID-19 would be limited to approximately 85,800. If we apply the CFR to *C*_*Phase*1_of the United States, we expect in excess of 12,300 fatalities resulting from COVID-19 by March 31.

Applying the same CFR to *C*_*Stabilized*_, we expect about 3,800 deaths. If the United States implements the measures noted in South Korea, Japan, and Iran, approximately 8,500 of deaths could be prevented.

The *Phase 2* nations we examined, Australia plus the developing nations of Brazil, Russia, India, and South Africa, have a unique opportunity to compare the differences between the stabilized nations and expanding *Phase 1* nations. India, the second most populous country in the world, has a population density over 3 times greater than that of China (454 vs 145 persons per square kilometer)^21^. This is far greater than the remaining *Phase 2* nations, indicating the greatest potential for COVID-19 expansion. If India expands as per *C*_*Phase*1_, we expect approximately 123,000 cases, resulting in over 6,800 fatalities, by April 25. However, if India expands as per *C*_*Stabilized*_, we expect 1,745 cases. Given the high attack rate and rapid spread of COVID-19 in densely populated, confined areas such as cruise ships, prisons, and hospitals, *I*_*2500*_ may underestimate India if its dense urban population interacts normally. However, if India successfully executes its 21-day lockdown on its 1.3 billion people, we calculate approximately 5,500 lives saved by April 16.

Our model may prove to be optimistic when considering the application of *Phase 1 Country*’s largely developed healthcare systems with those of the *Phase 2* developing countries. South Africa, Brazil, India, and have 2.8, 2.2, and 0.7 hospital beds per 1000 people, respectively.

Though closer to Iran’s 1.5 hospital beds per 1000 people, they are far below Japan and South Korea with 13.4 and 11.5, respectively^22^. Therefore, even with robust laboratory testing and quarantine measures, we must prepare for greater COVID-19 growth in developing countries. It is equally important to note that South Korea has a population density in excess of India (530 vs 455 people per square kilometer)^21^. Japan also exceeds the remainder of the studied nations with 347 people per square kilometer. Therefore, a country’s ability to manage COVID-19 is not primarily limited by population or population density.

Looking forward, we advise countries to rapidly implement policies to augment the spread of COVID-19. The exponential spread of disease is difficult to intuitively explain to the general population. In everyday life, we encounter arithmetic changes more often than exponential changes. Our instinct of examining relatively manageable changes to the case load to make policy decisions will quickly be eclipsed by the exponential expansion of this disease. Further complicating the matter is an estimation that 17.9% of COVID-19 patients are asymptomatic^23^. Looking at the data from the *Diamond Princess* cruise line, most of the cases occurred before or around the start of the quarantine. This reemphasizes the importance to make proactive, as opposed to reactive, policy decisions. It additionally demonstrates the importance of individual responsibility towards social distancing and personal hygiene. Should countries integrate the methods behind the success noted in South Korea, Japan, and Iran, thousands, perhaps millions, of deaths be prevented.

### Limitations

It is important to consider the significant limitations of this preliminary study. First, our model assumes continuous, regular exponential growth. Disease epidemics ultimately follow a sigmoidal shape, as they approach the carrying capacity of the disease. SEIR models account for a decreasing susceptible population and increasing recovered population with immunity. These models prove to be superior in long term analysis of a disease’s expansion. It is most likely that our model is only applicable for the initial exponential expansion of a disease. For COVID-19, we caution its usage beyond 60 days after initial seeding of 100 cases.

The data collected by the JHU CSSE is impacted by the reporting capabilities of each country. Subclinical cases are improbable to detect through surveillance screening due to lack of laboratory resources in most countries. Such cases would be entirely missed as presumptive cases, as these patients would not report to a healthcare facility. As mentioned earlier, the presumptive asymptomatic SARS-COV-2 positive population is 17.9%^23^. Beyond sub-clinical cases, governments are instructing mildly symptomatic patients to quarantine at home rather than seek hospital care. These cases would also avoid detection and registration in the data set.

The model equations serve to average the differences between healthcare systems, hospital bed per capita, laboratory testing capabilities, population dynamics, etc. When applying the model to any given country in initial phase of growth (<60 days after 100 cases), though, these differences are expected to exert considerable influence on COVID-19’s growth rate. This further emphasizes the caution to utilize this model beyond the initial expansion of the disease.

## Data Availability

Our study examines this pandemic through data provided by the Johns Hopkins University’s Systems Science and Engineering Group (JHU CSSE. It is an aggregation of case data starting on January 22, 2020
CSSEGISandData. 2019 Novel Coronavirus COVID-19 (2019-NCoV) Data Repository by Johns Hopkins CSSE. Johns Hopkins University Center for Systems Science and Engineering; 2020. https://github.com/CSSEGISandData/COVID-19. Accessed March 23, 2020.

